# Dissecting Treatment Response in Depression: A Symptom Network Analysis of rTMS Effects

**DOI:** 10.1101/2025.11.04.25339494

**Authors:** Luise Victoria Claaß, Marius Gruber, Delia Scheepers, Jonathan Repple, Nils Opel

**Author notes:** shared first authorship (Luise Victoria Claaß, Marius Gruber). shared last authorship (Jonathan Repple, Nils Opel).

## Abstract

Increasing evidence suggests that personalizing treatment for depression requires a refined understanding of how interventions affect distinct symptom domains, rather than focusing on total symptom scores. Different treatments may influence specific symptoms in unique ways, and these symptoms can interact dynamically - a relationship that can be explored using network modeling.

In this observational, naturalistic study, we applied single-symptom and network analyses to examine symptom changes in depressed inpatients (total n=108) receiving either treatment as usual (TAU; psychotherapy and pharmacotherapy) (n=62) or adjunctive repetitive transcranial magnetic stimulation (rTMS) (n=46). While overall symptom reduction and changes within individual symptom domains did not differ significantly between groups, network analyses revealed marked differences in symptom interrelations: In the TAU group, *loss of energy* and *depressed mood* emerged as the most central symptoms within the course of symptom change, whereas in the rTMS group, *loss of interest* and *decreased concentration* were most central and influential.

These findings highlight the importance of examining treatment effects at the single-symptom level and considering the complex interconnections among symptoms when evaluating and personalizing interventions for depression.

## Introduction

Major depressive disorder (MDD) is a heterogeneous condition characterized by diverse constellations of symptoms, including affective, cognitive, and somatic dimensions. Although current treatments - such as pharmacotherapy, psychotherapy, and neuromodulation - are generally effective on average, individual responses vary widely. Increasing evidence suggests that personalizing treatment for depression requires a refined understanding of how specific interventions affect distinct symptom domains rather than focusing solely on total symptom scores (Fried, 2017; Fried & Nesse, 2015; Fried & Robinaugh, 2020). Different treatments may influence particular symptoms in unique ways, and these symptoms interact dynamically within individuals over time. Network modeling offers a powerful framework to investigate these interrelations and their changes across treatments (McNally, 2016).

In network theory, phenomena are conceptualized as systems of interconnected components. Networks consist of nodes and edges, where nodes represent the objects of study and edges the connections between them. In psychopathology networks, nodes correspond to individual symptoms, and edges represent statistical associations between symptoms (Borsboom, 2017). Rather than assuming that disorders arise from a single latent cause, network analysis views mental disorders as systems of mutually reinforcing symptoms (e.g., insomnia increasing fatigue, which in turn intensifies concentration problems).

Within such networks, edge thickness indicates the strength of association between symptoms, and centrality metrics quantify the relative importance of each node. Several measures of centrality have been proposed (Freeman, 1978), among which strength centrality, which is the sum of the absolute weights of all edges connected to a node, is particularly relevant for psychopathology research. Highly central symptoms are those most strongly connected to others and may thus play key roles in the maintenance or remission of depressive states. For instance, activation of a highly central symptom is more likely to propagate through the network, potentially triggering other symptoms. Partial correlation networks provide an especially informative approach for such investigations, as they estimate the unique association between each pair of symptoms while controlling for all others. This allows researchers to delineate the direct interconnections that may reflect potential causal pathways within the symptom network (Borsboom & Cramer, 2013).

Empirical evidence supports the clinical relevance of network properties. Van Borkulo et al. (2015) showed that individuals whose depression persisted over a two-year period exhibited more densely connected baseline symptom networks than those who remitted, with guilt, fatigue, and loss of energy emerging as especially central symptoms. Similarly, Fried et al. (2015) demonstrated that symptoms with high closeness centrality (those most efficiently connected to others) were particularly responsive to antidepressant medication. These findings suggest that network architecture may help identify treatment targets and predict recovery trajectories.

Repetitive transcranial magnetic stimulation (rTMS) represents one of the most rapidly evolving treatment options for depression, with recent studies demonstrating robust antidepressant effects on overall symptom reduction (Blumberger et al., 2018; Cole et al., 2020). In contrast to pharmacotherapy and psychotherapy - as well as to other established neuromodulatory interventions such as electroconvulsive therapy (ECT) - modern rTMS protocols offer the possibility to selectively target specific cortical regions. Current rTMS protocols for depression, such as intermittent theta-burst stimulation (iTBS) of the left dorsolateral prefrontal cortex (DLPFC), are thought to exert their therapeutic effects by modulating functional connectivity within distributed neural networks (Cash et al., 2019; Weigand et al., 2018). Considering its unique mechanistic profile, rTMS may exert its antidepressant effects by modulating distinct symptom domains rather than producing uniform improvements across all symptoms. However, most clinical studies to date have evaluated rTMS outcomes using global depression sum scores, without accounting for potential heterogeneity across symptom dimensions. Consequently, it remains unclear whether - and in which specific symptom domains - rTMS produces differential effects compared to established standard treatments.

Building on this framework, the present study applied network analysis to examine how adjunctive rTMS for depression influences the interrelations among individual depressive symptoms, compared with standard inpatient treatment alone, typically consisting of pharmacotherapy and psychotherapy. By modeling symptom changes between baseline and follow-up, we aim to elucidate whether and how these interventions differentially alter the structure and centrality of symptom networks. Understanding treatment-specific patterns of symptom interconnectivity may provide novel insights into symptom-specific patterns of treatment response and inform more individualized treatment strategies for depression.

## Methods

### Study Procedure

This study was conducted as an observational longitudinal investigation involving in-patients recruited at the university hospitals in Jena and Frankfurt am Main from 2022-2025. Participants were screened and enrolled during their stay on psychiatric in-patient wards by study personnel. Eligible participants were patients with a diagnosis of either major depressive disorder (MDD) or bipolar disorder (BD), confirmed through a structured clinical interview conducted by trained study personnel according to the criteria of the DSM-5.

All participants received standard antidepressant treatment consisting of combined pharmacotherapy and psychotherapy as determined by their treating clinicians within the framework of routine clinical care. In addition, a subset of patients underwent adjunctive repetitive transcranial magnetic stimulation (rTMS) when this treatment modality was considered clinically appropriate and was recommended by the medical staff responsible for patient care.

For both study sites (Jena and Frankfurt), rTMS was administered using intermittent theta-burst stimulation (iTBS) targeting the left dorsolateral prefrontal cortex (DLPFC), localized with the Beam F3 method (Beam et al., 2009). Stimulation intensity was set at a minimum of 80% of the individual resting motor threshold. In Jena, participants received a total of 30 treatment sessions delivered over 15 days (two sessions per day), consisting of bilateral stimulation with iTBS over the left DLPFC and continuous TBS (cTBS) over the right DLPFC. The Frankfurt protocol comprised 20-30 sessions delivered over 6 weeks (one session per day), with unilateral iTBS targeting the left DLPFC.

Data were collected at two time points. The first assessment took place at the beginning of the in-patient treatment period, or - if rTMS treatment was planned - before the first rTMS session. The second assessment occurred after the completion of the rTMS treatment or, in patients who did not receive rTMS, on average 6 weeks after beginning inpatient treatment. This design allowed for the longitudinal observation of clinical and biological changes under naturalistic treatment conditions.

At baseline, during treatment, and throughout the follow-up period, questionnaires were administered by study personnel.

### Participants

A total of 104 participants were included from the Universities of Jena and Frankfurt am Main. Patients were recruited between October 2022 and August 2025. Participants were eligible for inclusion if they were between 18 and 75 years of age and had a clinically confirmed diagnosis of an affective disorder, including major depressive disorder or bipolar disorder.

Exclusion criteria comprised the presence of substance dependence or any disorder within the psychotic spectrum. Since participants of this study also underwent magnetic resonance imaging (MRI) assessments, individuals with contraindications to MRI scanning, such as non–MRI-compatible prostheses, metal implants, or other metallic materials, were excluded. Further exclusion criteria included any neurological disorder affecting the brain or nervous system, such as migraine with aura (whereas migraine without aura was not considered exclusionary), encephalopathy, narcolepsy, neuroborreliosis, stroke, meningitis, multiple sclerosis, Parkinson’s disease, Alzheimer’s disease, paralysis, dementia, or neuronal muscle atrophy. Participants with autoimmune or systemic diseases were also excluded, as well as those with current or previous malignant disease, except in cases of benign tumors located outside the brain. Additional exclusion criteria included moderate to severe hearing impairment and the current use of benzodiazepines. Ethical approval was granted by the institutional review boards of both participating centers, and all participants provided written informed consent in accordance with the Declaration of Helsinki.

To maximize statistical power and enhance the informativeness of the analysis, all patients enrolled up to August 2025 were included in the present analysis. Descriptives of the cohort can be found in Table 1.

**Table 1.** Sociodemographic and clinical characteristics of the sample. * t- or X^2^-tests.

| **Variable** | **TAU**<br>**Mean (SD) or %** | **rTMS**<br>**Mean (SD) or %** | **p\*** |
| --- | --- | --- | --- |
| n | 62 | 46 |  |
| Female (%) | 54.8% | 50% | 0.762 |
| Age (years) | 33.11 (12.81) | 46.67 (14.55) | <0.001 |
| BDI baseline (sum) | 31.58 (10.63) | 30.28 (9.58) | 0.508 |
| HAM-D baseline (sum) | 17.15 (8.34) | 17.10 (8.01) | 0.977 |

### Interventions

#### rTMS

The resting motor threshold was determined at the beginning of rTMS treatment and reassessed as needed. It was defined as the minimal stimulation intensity that evoked a visible contraction in the muscle of the right thumb. The Beam F3 method was used to localize the left and right dorsolateral prefrontal cortex (DLPFC, Beam et al., 2009). rTMS was administered using an iTBS protocol over the left DLPFC, consisting of triplet 50 Hz bursts repeated at 5 Hz, with 2 seconds on and 8 seconds off, totaling 600 pulses per session with a duration of 3 minutes and 9 seconds. The intensity was selected in increments from 80% to 120% of the resting motor threshold. For Jena participants over the right DLPFC, a cTBS protocol was used, delivering 1800 pulses in a continuous train of 600 theta bursts lasting 40 seconds. Patients were free to discontinue the treatment at any time in case of side-effects or for any other reason.

#### Treatment as usual

Standard inpatient treatment (TAU) consisted of combined treatment with pharmacotherapy and psychotherapy at both sites. With regard to psychotherapy, all patients received sessions of cognitive-behavioral therapy, at least once a week. Medication consisted of pharmacological treatment steps following the protocol in the medication algorithm of the German guidelines (BAEK, KBV, AMWF, 2022). The next treatment step consisted of either a switch from the current antidepressant medication to a tricyclic antidepressant or augmentation of the current antidepressant medication with lithium or a second-generation antipsychotic. If this was not a suitable option (e.g., for medical reasons), a different antidepressant could be prescribed, as would be the case during usual care.

#### Measures

The Hamilton Depression Rating Scale (HAM-D) and Beck Depression Inventory (BDI-II) were used to assess severity of depressive symptoms at baseline and during treatment (Hamilton, 1960; Beck et al., 1996; Wang & Gorenstein, 2013). To examine the effect of rTMS versus TAU on depressive symptom dimensions, 9 items of the BDI-II were chosen, which represented most closely the 9 DSM-V criteria for MDD. An overview of these items can be found in Table 2. The BDI-II was selected for this study due to its strong psychometric properties, including high reliability, sensitivity, and specificity, as well as its well-established concurrent and content validity (Wang & Gorenstein, 2013; Storch et al., 2004). Moreover, the BDI-II is one of the most widely used self-report instruments for assessing depressive symptoms in both research and clinical practice, ensuring that our findings are directly translatable to real-world clinical settings and comparable across studies.

**Table 2.** BDI-II items corresponding DSM-V criteria for MDD.

| **Abbreviation** | **Symptom** | **BDI-II item** |
| --- | --- | --- |
| Dm | Depressed Mood | BDI 1 |
| Li | Loss of interest | BDI 12 |
| Ap | Change of appetite | BDI 18 |
| SI | Insomnia / Hypersomnia | BDI 16 |
| Pa | Psychomotor agitation | BDI 11 |
| Le | Loss of energy | BDI 20 |
| Fw | Feeling worthless | BDI 3 |
| Dc | Decreased concentration | BDI 19 |
| St | Thoughts of death / suicide | BDI 9 |

### Statistical Analysis

#### Handling of missing data

If less than 5% of data was missing, BDI-II items were imputed using predictive mean matching. Changes in total scores of the BDI-II and HAM-D were then analyzed using a repeated-measures analysis of variance (ANOVA) with Time (T0, T1) as a within-subject factor and Treatment (TAU, rTMS) as a between-subject factor.

Second, in order to capture the core clinical features that characterize a diagnosis of MDD in a manner that closely reflects the clinical context and ensures reproducibility, we selected nine items from the BDI-II corresponding to DSM-5 diagnostic criteria for MDD (Table 2). For this item-level analyses, each of the nine depressive symptoms (depressed mood, loss of interest, appetite change, sleep disturbance, psychomotor agitation, loss of energy, feelings of worthlessness, decreased concentration, and suicidal ideation) was treated as a separate dependent variable. A repeated-measures ANOVA was performed with Time and Symptom as within-subject factors and Treatment as the between-subject factor. The critical interactions Time × Treatment and Time × Symptom × Treatment tested whether pre–post changes differed between groups and whether these differences varied across individual symptoms.

All analyses were conducted in R (v4.4.3) using the *afex* package (Singmann, 2024). Statistical significance was set at *p* < 0.05 (two-tailed).

### Network estimation

To examine symptom interrelations and their change following treatment, we estimated separate partial correlation networks for patients receiving TAU and those undergoing rTMS.

For each participant, symptom change over time was calculated as the difference between baseline (T0) and follow-up (T1) scores for the nine selected BDI-II items, yielding Δ-scores defined as T0 minus T1 for each item. This approach ensures that positive Δ-scores represent symptom improvement, consistent with the interpretation of BDI-II where higher values indicate greater severity. The resulting dataset consisted of nine Δ-score nodes for the core depressive symptoms: Depressed Mood, Loss of Interest, Change of Appetite, Change of Sleep, Psychomotor Agitation, Loss of Energy, Feelings of Worthlessness, Decreased Concentration, and Suicidal Thoughts.

Prior to network estimation, all variables were assessed for near-zero variance and collinearity to ensure the integrity of correlation calculations. The network structure was then estimated using the estimateNetwork() function from the *bootnet* package (Epskamp et al., 2018), with the default model set to “pcor” to compute partial correlation networks. In this framework, each edge represents a unique association between two nodes after controlling for all other variables in the network. Partial correlations were estimated as Spearman’s rank correlations to enhance stability and robustness, particularly in the context of ordinal variables and limited sample sizes, addressing potential issues of non-positive definite matrices and category sparsity. Resulting networks were visualized using *qgraph* (Epskamp et al., 2012), with edge thickness and color (positive correlations depicted as green edges, negative ones as red) reflecting the magnitude and direction of partial correlations.

### Centrality and Stability Analyses

Node-level centrality indices (Strength, Closeness, Betweenness) were computed to quantify each symptom’s relative influence within the network structure.

To assess the robustness and reliability of network characteristics, case-dropping bootstrapping was conducted. For each group, 1,000 bootstrap samples were drawn while progressively reducing the number of cases, enabling calculation of the Correlation Stability (CS) coefficient for centrality indices. The CS coefficient quantifies how much of the sample can be dropped while maintaining a correlation of ≥ 0.7 between original and resampled centrality estimates (Epskamp et al., 2018). Stability analyses focused on node strength, reflecting each symptom’s degree of connectivity within the network. Bootstrapped confidence intervals and stability plots were visually inspected to evaluate the robustness of the estimated network structures.

## Results

### Participants

Overall, 108 patients were recruited at both sites. TAU and rTMS groups did not differ in sex or depression severity. Age differed significantly, with the rTMS group showing an older age distribution. The average time between baseline and follow-up was 45.6 days (SD 14.4). Of all 108 patients included, 12 had a bipolar diagnosis and were currently in a depressive episode, while the rest were diagnosed with MDD. The clinical characteristics are presented in Table 1.

### Treatment Outcomes

Both groups showed a marked improvement in overall depression sum scores in both the BDI-II and HAM-D. For 10 subjects, no sum scores for the HAM-D at T1 were available.

A repeated-measures ANOVA with the within-subject factor Time (T0, T1) and the between-subject factor Treatment (TAU, rTMS) revealed a significant main effect of Time, indicating an overall reduction in depressive symptoms from pre-to post-treatment (F(1,96) = 109.82, *p* < 0.001). The main effect of Treatment was not significant, suggesting comparable baseline severity between groups. Importantly, the Time × Treatment interaction was not significant, indicating that the degree of symptom improvement did not differ between the two treatment conditions (F(1,96) = 0.39, *p* = 0.535; Figure 1).

**Figure 1.**
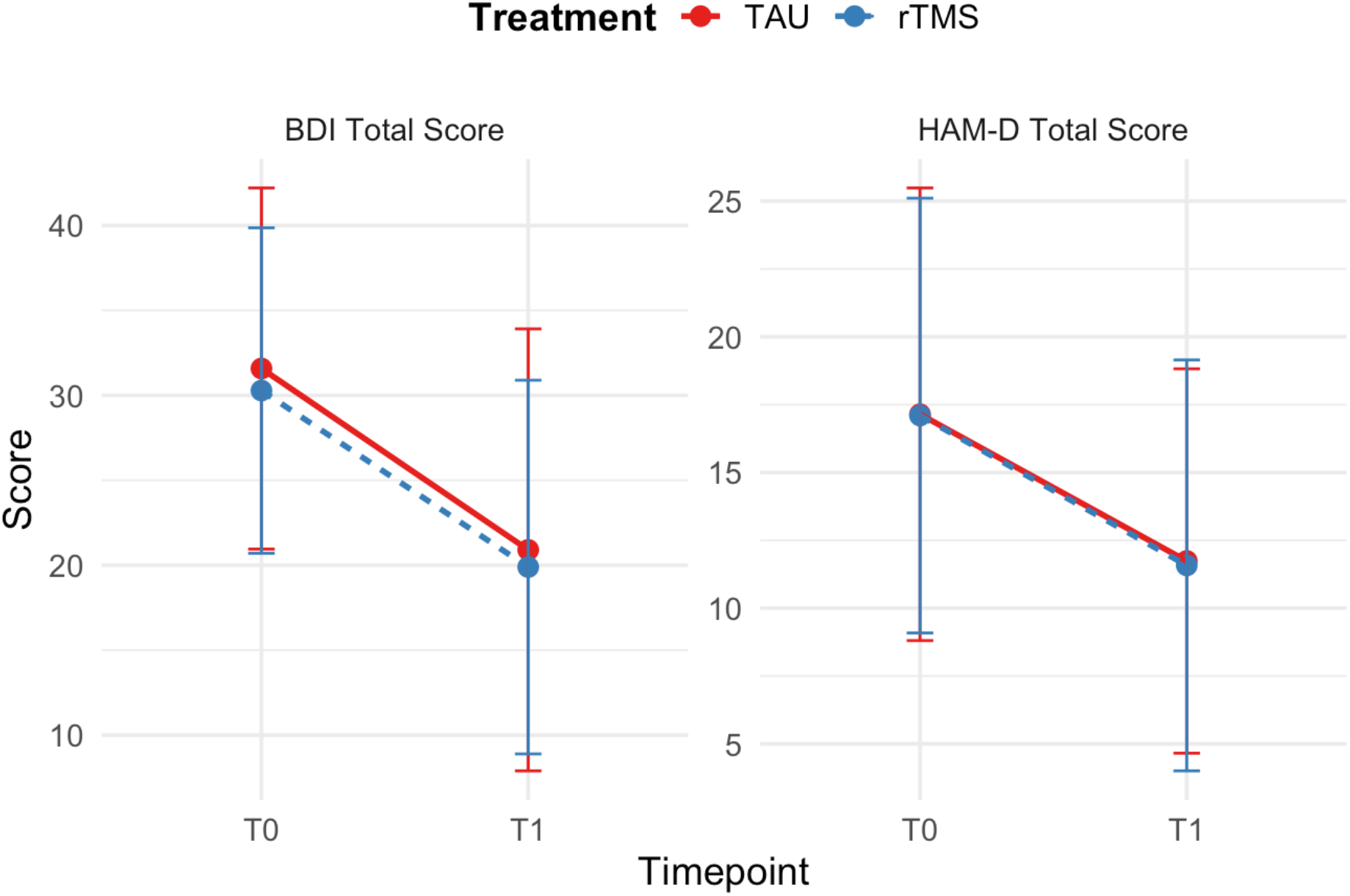
Change in depressive symptom scores from baseline (T0) to follow-up (T1). Results are presented separately for the treatment as usual (TAU; red) and repetitive transcranial magnetic stimulation (rTMS; blue) groups. BDI = Beck Depression Inventory (BDI-II); HAMD = Hamilton Depression Rating Scale (HAM-D).

When examining the change in the 9 BDI-II items that represent DSM-V diagnostic criteria (Table 2), both treatment groups showed reductions in depressive symptoms across all items, indicating an improvement in the respective depressive symptoms (Figure 2). In the repeated measures ANOVA a significant main effect of Time was observed (F(1,106) = 78.13, *p* < 0.001), whereas there was no significant Time x Treatment (F(1,106) = 0.25, p = 0.619) or Time x Treatment x Symptom interaction (F(8,848) = 0.64, p = 0.740), indicating there were no notable differences in symptom change, neither between the two groups, nor between items.

**Figure 2.**
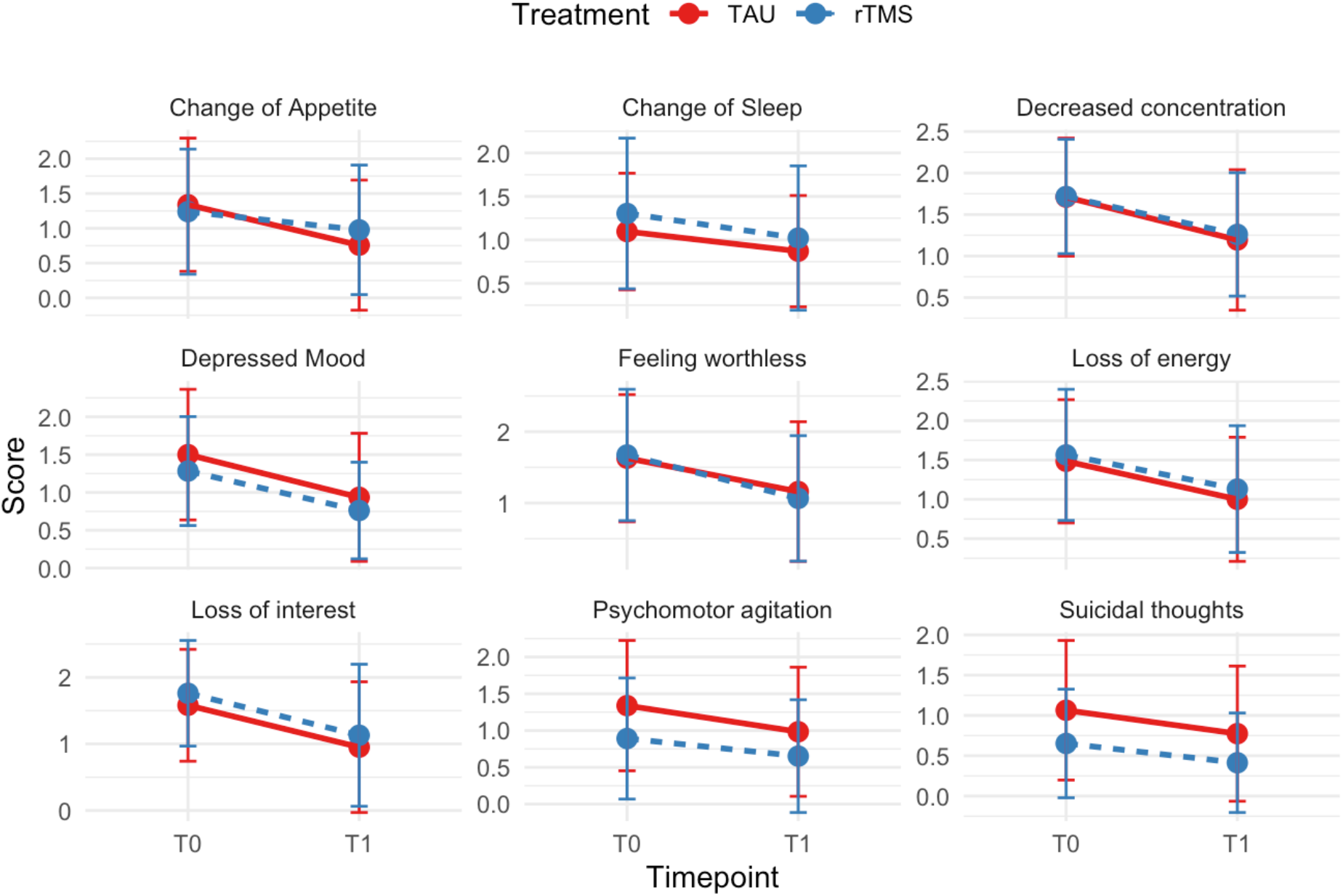
Change in BDI-II item scores from baseline (T0) to follow-up (T1). The assignment of symptoms shown in the figure to BDI-II items is detailed in Table 2. Results are presented separately for the treatment as usual (TAU; red) and repetitive transcranial magnetic stimulation (rTMS; blue) groups.

### Network analysis

Partial correlation networks, estimated separately for each treatment group, with 9 nodes representing Δ-scores defined as BDI-II items at T0 minus T1 (i.e., positive Δ-scores indicating symptom improvement), revealed significant differences in how symptom score changes relate to each other. Visual inspection of plots indicated that overall, the number and strength of connections were higher in the rTMS group compared to the TAU network (Figure 3). Comparing the different associations among symptoms, in the TAU group, the strongest connection was between Depressed mood (Dm, Figure 3, left panel) and Suicidal thoughts (St). Slightly less strong were the connections between Depressed mood and Loss of interest (Li) and between Suicidal thoughts and Feeling worthless (Fw).

**Figure 3.**
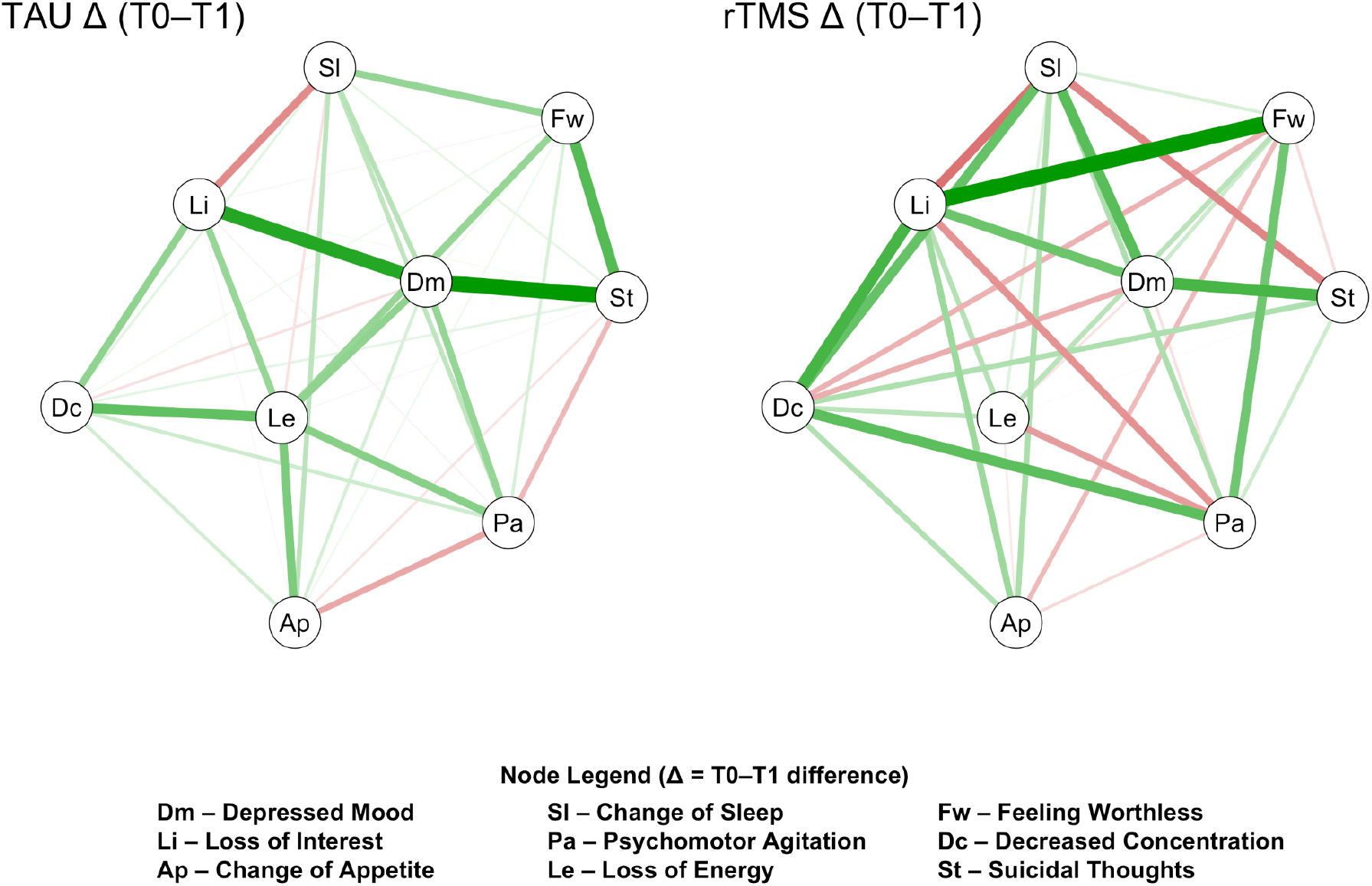
Partial correlation networks of depressive symptom change in the TAU and rTMS groups. Partial correlation networks estimated using Spearman correlations are shown separately for patients receiving treatment as usual (TAU; left) and repetitive transcranial magnetic stimulation (rTMS; right). Each node represents the change score (Δ = T0-T1) of a depressive symptom item from the Beck Depression Inventory-II (BDI-II), where positive Δ values indicate symptom improvement over time. Edges represent regularized partial correlations between symptom changes, controlling for all other nodes in the network. Thicker edges indicate stronger associations, with green edges representing positive and red edges representing negative partial correlations. Node labels correspond to specific BDI-II items, as detailed in the legend (bottom panel).

In the rTMS network, the strongest connection was observed between Loss of interest and Feeling worthless. Other prominent edges include those between Decreased concentration (Dc) and Loss of interest, and between Decreased concentration and Psychomotor agitation (Pa). Depressed mood showed numerous strong edges with other items, such as Loss of interest, Change of Sleep, and Suicidal thoughts.

Figure 4 shows the centrality indices: strength (the sum of absolute edge weights connected to a node), betweenness (how much a node lies on the shortest paths between other nodes), and closeness (the inverse of the average distance from a node to all others). Higher values indicate a greater influence of the symptom within the network. Notably, while in the TAU network, depressed mood and loss of energy exhibit the highest values for all three centrality indices and are therefore well-connected within the network, they play a minor role in the rTMS network, showing only small to moderate values for strength, betweenness, and closeness. In comparison, loss of interest and decreased concentration exhibited higher centrality indices in this correlation network.

**Figure 4.**
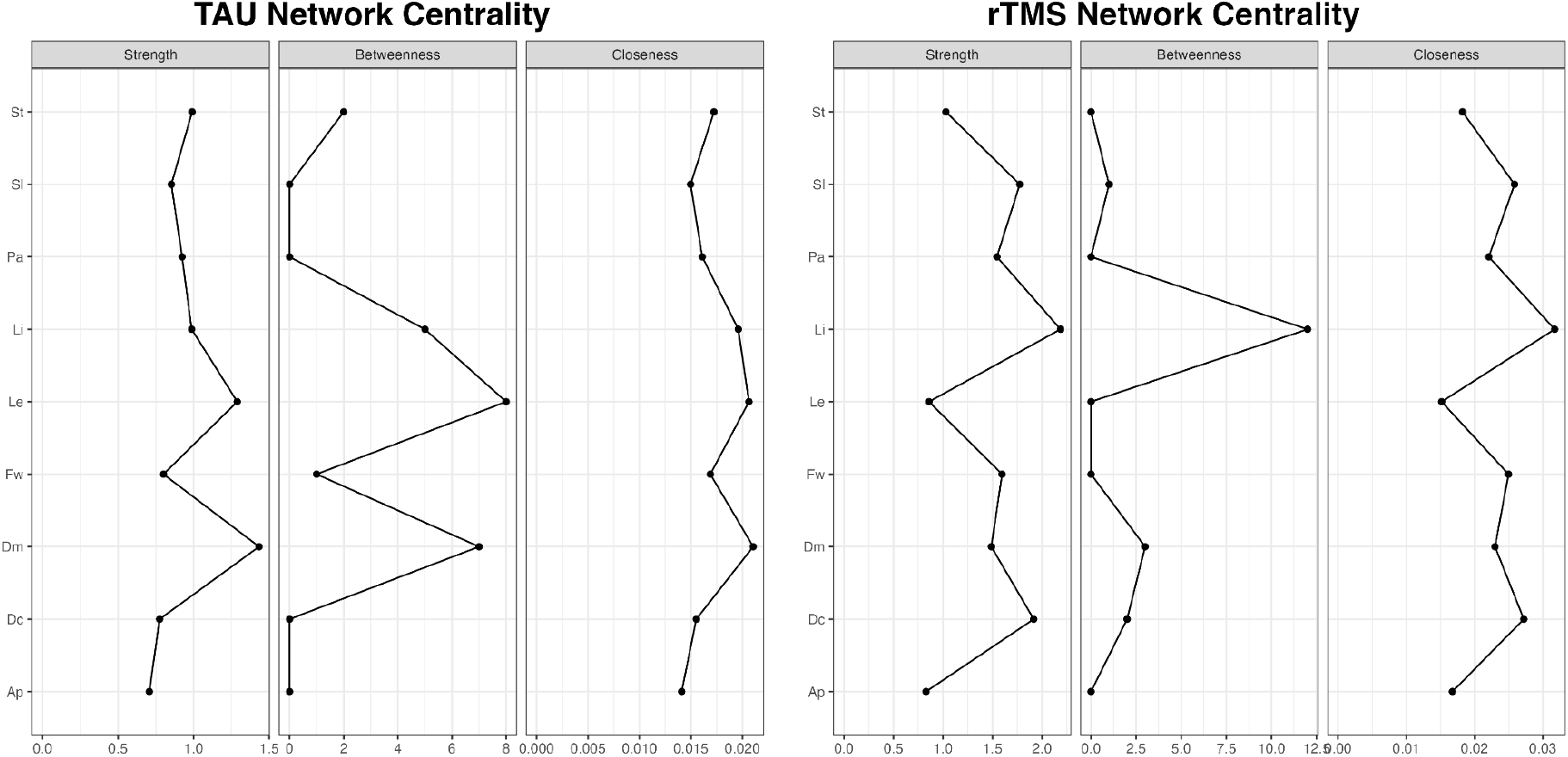
Centrality indices of depressive symptom networks in the TAU and rTMS groups. Centrality plots display the relative importance of each depressive symptom within the partial correlation networks for patients receiving treatment as usual (TAU; left) and repetitive transcranial magnetic stimulation (rTMS; right). Node centrality was quantified using three standard indices: strength (sum of absolute edge weights connected to a node), betweenness (extent to which a node lies on the shortest paths between other nodes), and closeness (inverse of the average distance from a node to all others). Higher values indicate greater influence of the corresponding symptom within the network. Node labels correspond to the Beck Depression Inventory-II (BDI-II) items representing DSM-5 depressive symptoms.

### Stability analysis

Case-dropping bootstrapping with 1,000 bootstrap samples revealed Correlation Stability (CS) coefficients of 0.05 for the TAU group and 0.196 for the rTMS group. This is lower than the average CS coefficient values deemed stable, according to established guidelines being 0.25 (Epskamp et al., 2018). Visual inspection of the bootstrapped strength centrality plot revealed a large overlap of 95%-confidence intervals, although Loss of interest and Decreased concentration were the items where CI overlap was the least (Figure 5).

**Figure 5.**
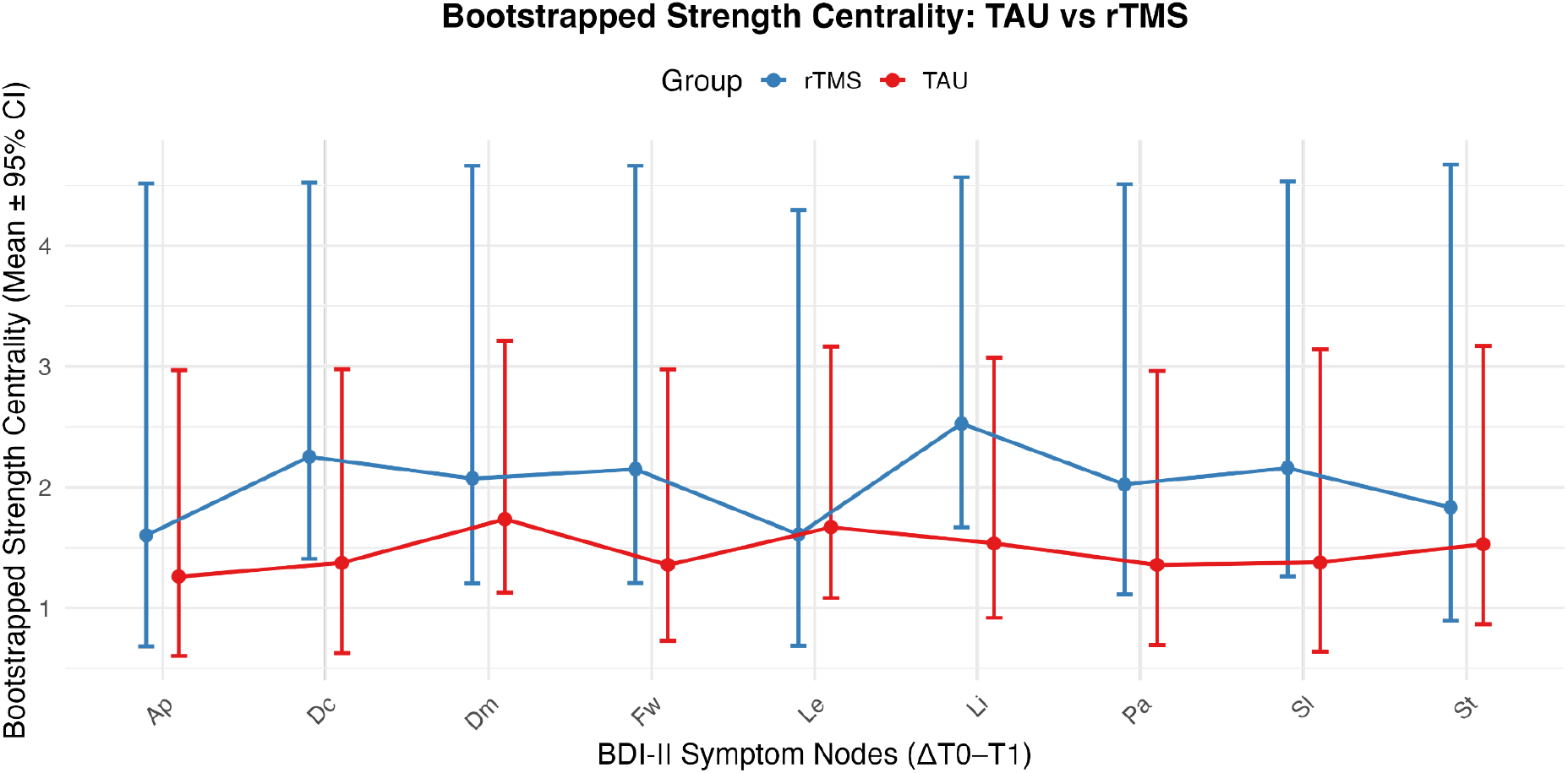
Bootstrapped strength centrality of depressive symptom networks in the TAU and rTMS groups. Mean bootstrapped strength centrality estimates (±95% confidence intervals) are shown for each Beck Depression Inventory-II (BDI-II) symptom node, representing the change scores (ΔT0-T1). Results are presented separately for the treatment as usual (TAU; red) and repetitive transcranial magnetic stimulation (rTMS; blue) groups. Strength centrality reflects the overall connectedness of each symptom within the network, with higher values indicating greater influence on the overall symptom structure.

## Discussion

In the present study, we demonstrate differential patterns of symptom network change following rTMS compared with standard inpatient treatment. While overall depressive symptom severity and changes in individual symptom scores did not differ significantly between groups, the interrelations among symptom change - captured through network analysis - showed distinct patterns. These findings indicate that both interventions may influence the structure of symptom change differently at the network level.

Specifically, *depressed mood* and *loss of energy* emerged as the most central symptoms in the TAU group, whereas *loss of interest* and *decreased concentration* were most central in the rTMS network. This suggests that rTMS and standard treatment may engage different pathways of symptom improvement, with certain symptom domains acting as key mediators of recovery within each intervention.

These findings carry potential implications for personalized psychiatry. The differentially central symptoms observed across the TAU and rTMS networks may serve as targets for future individualized treatment strategies. For instance, if loss of interest or decreased concentration consistently emerge as highly central in response to rTMS, these symptoms could inform treatment selection or parameter optimization. Nevertheless, such conclusions remain preliminary and require replication and validation in larger, independent samples. Conceptually, precision psychiatry could integrate pre-treatment symptom network analyses to identify the most central symptoms for each individual, thereby enabling clinicians to tailor interventions to a patient’s unique symptom architecture.

Overall, our results highlight the potential of network analysis to capture nuanced, symptom-specific treatment effects that extend beyond conventional measures such as total scores or single-symptom changes. Future studies should replicate and extend this approach using longitudinal data with more fine-grained temporal resolution to better capture the dynamic interplay of symptoms over the course of treatment. Elucidating how specific interventions modify symptom networks over time could ultimately form the basis for more personalized and targeted treatment strategies tailored to distinct clinical phenotypes.

Several limitations should be acknowledged. First, differences in treatment protocols between sites - such as variations in stimulation parameters and treatment intensity - may have influenced the observed network structures and limit direct comparability. Second, the relatively small sample size reduces statistical power and may restrict the generalizability of the findings. Third, the absence of blinding and randomization introduces the possibility of expectancy and site effects, which cannot be fully ruled out. Additionally, patient characteristics that led to the specific treatment decision from a clinical perspective might have an influence on baseline symptom networks and therefore all subsequent analyses. Finally, the study design focused on pre- and post-treatment comparisons; future work incorporating repeated, fine-grained longitudinal assessments could provide deeper insight into the temporal evolution of symptom networks and causal directions of change.

In sum, these findings support the utility of network-based approaches for capturing the complex, multidimensional nature of treatment response in depression. By revealing how distinct interventions -such as rTMS and multimodal inpatient therapy - differentially reshape symptom interrelations, this framework moves beyond aggregate symptom scores toward a more mechanistic understanding of clinical change. Continued integration of symptom network analysis with neurobiological and behavioral markers may ultimately facilitate the development of personalized, circuit-targeted treatment strategies for mood disorders.

## Data Availability

All data produced in the present study are available upon reasonable request to the authors.

## References

Beam, W., Borckardt, J. J., Reeves, S. T., & George, M. S. (2009, Jan). An efficient and accurate new method for locating the F3 position for prefrontal TMS applications. Brain Stimul, 2(1), 50–54. 10.1016/j.brs.2008.09.006

Beck, A. T., Steer, R. A., & Brown, G. (1996). Beck depression inventory–II. Psychological assessment.

Blumberger, D. M., Vila-Rodriguez, F., Thorpe, K. E., Feffer, K., Noda, Y., Giacobbe, P., Knyahnytska, Y., Kennedy, S. H., Lam, R. W., Daskalakis, Z. J., & Downar, J. (2018, Apr 28). Effectiveness of theta burst versus high-frequency repetitive transcranial magnetic stimulation in patients with depression (THREE-D): a randomised non-inferiority trial. Lancet, 391(10131), 1683–1692. 10.1016/s0140-6736(18)30295-2

Borsboom, D. (2017, Feb). A network theory of mental disorders. World Psychiatry, 16(1), 5–13. 10.1002/wps.20375

Borsboom, D., & Cramer, A. O. (2013). Network analysis: an integrative approach to the structure of psychopathology. Annu Rev Clin Psychol, 9, 91–121. 10.1146/annurev-clinpsy-050212-185608

Cash, R. F. H., Zalesky, A., Thomson, R. H., Tian, Y., Cocchi, L., & Fitzgerald, P. B. (2019, Jul 15). Subgenual Functional Connectivity Predicts Antidepressant Treatment Response to Transcranial Magnetic Stimulation: Independent Validation and Evaluation of Personalization. Biol Psychiatry, 86(2), e5-e7. 10.1016/j.biopsych.2018.12.002

Cole, E. J., Stimpson, K. H., Bentzley, B. S., Gulser, M., Cherian, K., Tischler, C., Nejad, R., Pankow, H., Choi, E., Aaron, H., Espil, F. M., Pannu, J., Xiao, X., Duvio, D., Solvason, H. B., Hawkins, J., Guerra, A., Jo, B., Raj, K. S., Phillips, A. L., Barmak, F., Bishop, J. H., Coetzee, J. P., DeBattista, C., Keller, J., Schatzberg, A. F., Sudheimer, K. D., & Williams, N. R. (2020, Aug 1). Stanford Accelerated Intelligent Neuromodulation Therapy for Treatment-Resistant Depression. Am J Psychiatry, 177(8), 716–726. 10.1176/appi.ajp.2019.19070720

Epskamp, S., Borsboom, D., & Fried, E. I. (2018, Feb). Estimating psychological networks and their accuracy: A tutorial paper. Behav Res Methods, 50(1), 195–212. 10.3758/s13428-017-0862-1

Epskamp, S., Cramer, A. O. J., Waldorp, L. J., Schmittmann, V. D., & Borsboom, D. (2012, 05/24). qgraph: Network Visualizations of Relationships in Psychometric Data. Journal of Statistical Software, 48(4), 1–18. 10.18637/jss.v048.i04

Freeman, L. C. (1978, 1978/01/01/). Centrality in social networks conceptual clarification. Social Networks, 1(3), 215–239. 10.1016/0378-8733(78)90021-7

Fried, E. I. (2017, Jan 15). The 52 symptoms of major depression: Lack of content overlap among seven common depression scales. J Affect Disord, 208, 191–197. 10.1016/j.jad.2016.10.019

Fried, E. I., Boschloo, L., van Borkulo, C. D., Schoevers, R. A., Romeijn, J.-W., Wichers, M., de Jonge, P., Nesse, R. M., Tuerlinckx, F., & Borsboom, D. (2015, 2015-August-21). Commentary: “Consistent Superiority of Selective Serotonin Reuptake Inhibitors Over Placebo in Reducing Depressed Mood in Patients with Major Depression” [General Commentary]. Frontiers in Psychiatry, Volume 6 - 2015. 10.3389/fpsyt.2015.00117

Fried, E. I., & Nesse, R. M. (2015, Feb 1). Depression is not a consistent syndrome: An investigation of unique symptom patterns in the STAR*D study. J Affect Disord, 172, 96–102. 10.1016/j.jad.2014.10.010

Fried, E. I., & Robinaugh, D. J. (2020, Jul 14). Systems all the way down: embracing complexity in mental health research. BMC Med, 18(1), 205. 10.1186/s12916-020-01668-w

Hamilton, M. (1960, Feb). A rating scale for depression. J Neurol Neurosurg Psychiatry, 23(1), 56–62. 10.1136/jnnp.23.1.56

McNally, R. J. (2016, Nov). Can network analysis transform psychopathology? Behav Res Ther, 86, 95–104. 10.1016/j.brat.2016.06.006

Singmann, H. B., Bolker, B., Westfall, J., Aust, F., & Ben-Shachar, M. S. (2024). afex: Analysis of Factorial Experiments. R package version 1. 4-1. https://CRAN.R-project.org/package=afex

Storch, E. A., Roberti, J. W., & Roth, D. A. (2004). Factor structure, concurrent validity, and internal consistency of the Beck Depression Inventory-Second Edition in a sample of college students. Depress Anxiety, 19(3), 187–189. 10.1002/da.20002

van Borkulo, C., Boschloo, L., Borsboom, D., Penninx, B. W. J. H., Waldorp, L. J., & Schoevers, R. A. (2015). Association of Symptom Network Structure With the Course of Depression. JAMA Psychiatry, 72(12), 1219–1226. 10.1001/jamapsychiatry.2015.2079

Wang, Y. P., & Gorenstein, C. (2013, Oct-Dec). Psychometric properties of the Beck Depression Inventory-II: a comprehensive review. Braz J Psychiatry, 35(4), 416–431. 10.1590/1516-4446-2012-1048

Weigand, A., Horn, A., Caballero, R., Cooke, D., Stern, A. P., Taylor, S. F., Press, D., Pascual-Leone, A., & Fox, M. D. (2018, Jul 1). Prospective Validation That Subgenual Connectivity Predicts Antidepressant Efficacy of Transcranial Magnetic Stimulation Sites. Biol Psychiatry, 84(1), 28–37. 10.1016/j.biopsych.2017.10.028

